# Challenges in the distribution of antimicrobial medications in community dispensaries in Accra, Ghana

**DOI:** 10.1101/2023.01.31.23285234

**Authors:** Hannah Camille Greene, Kinga Makovi, Rafiatu Abdul-Mumin, Akhil Bansal, Jemima Frimpong

## Abstract

**Introduction:** Community distribution of medications in low- and middle-income countries has been shown to accelerate the emergence of antimicrobial resistance. The distribution of medications is often carried out by private vendors operating under constrained conditions. Yet patterns in medicine distribution—and their consequences—are not well understood. The aim of this study was to illuminate the challenges reported by employees of chemical shops and pharmacies throughout Accra. Our objectives are twofold: to 1) assess obstacles and challenges faced by medicine vendors during their sales of antibiotic and antimalarial medications, and 2) identify opportunities for improving community-level stewardship of antimicrobials.

**Methods:** Responses to open-ended questions from a survey of 80 shopkeepers in pharmacies and chemical shops throughout Accra were analyzed using the socioecological model of public health.

**Results:** Overall, shopkeepers most often reported constraints at the interpersonal and community levels of the socioecological model of public health. These included the prohibitive costs of medicines, customer attitudes, and customers’ attempts at self-medication and uninformed antimicrobial use. Other challenges included a lack of diagnostic testing, supply chain issues, and the larger economic and healthcare situation of the community.

**Discussion:** The safe and effective distribution of medications was truncated by three main sources of obstacles: financial insecurity among customers, challenges directly in the treatment of illnesses, and broader issues with the fragmented healthcare infrastructure affecting shopkeepers’ roles as health educators and gatekeepers of medicines.

**Conclusion:** These context-specific findings identify tractable challenges faced by medicine vendors in Ghana, with relevance to antimicrobial stewardship across resource-poor settings globally. Addressing barriers faced by shopkeepers would provide an opportunity for significantly improving the provision of medications, and ultimately healthcare, at the community level. Such efforts will likely expand access to populations who may otherwise be unable to access services in formal institutions of care.

## Introduction

Antibiotics and antimalarials are critically important life-saving medications, but their misuse and overuse are accelerating the emergence of antimicrobial resistance (AMR) [1–3], which is projected to cause 10 million annual deaths by 2050 [4]. Major drivers of AMR take place at the community level beyond the purview of hospitals, in the forms of over-distribution and inappropriate use [5, 6]. If a patient uses an incomplete or substandard antibiotic treatment course, selection pressure allows resilient microbes to persist with genetic mutations conferring resistance to an active ingredient [7–9]. Drug-resistant pathogens thus replicate and can be transmitted to others [10]. If community transmission occurs—and it does so at particularly rapid rates in settings without adequate sanitation [1, 11, 12]—the original medication is rendered ineffective [13].

The prudent use of antibiotic and antimalarial medications is crucial for slowing the emergence of resistant strains by reducing opportunities for the evolution of resistance [5]. Judicious use is better-defined for formal institutions of healthcare given the already established practice of restricting or adjusting the use of antibiotics through diagnostic testing and clinical observation, but best practices for stewardship at the community level are murkier [14]. Pharmacies and chemical shops in low- and middle-income countries play a key part in how medications and medical advice are accessed and serve as gatekeepers of medications [15]. Few studies globally [16–21] and fewer within Ghana [22–25] have studied the challenges in community pharmacy for antimicrobials. Therefore, the aim of this study is to assess the context-specific attitudes and challenges faced by medicine distributors in Accra, Ghana, in their sales of antibiotic and antimalarial medications.

Many neglected tropical diseases (NTDs) persist in Accra [26], with endemic malaria comprising up to 38% of outpatient and hospital cases [27]. However, uncomplicated malaria can be resolved with a three-day treatment course of antimicrobials accessible at over-the-counter vendors, and many NTDs can be treated with antibiotics. Drug-resistant malaria and bacterial pathogens have already been identified in Ghana [28–34] and the distribution of antimicrobials has yet to be comprehensively studied.

Approximately 80% of the Ghanaian workforce is employed in the informal private sector [35], characterized by sporadic and low wages. When healthcare is needed, the up-front cost of formal medical institutions can be prohibitive where a majority of the population faces high income insecurity [36]. Health finance primarily relies on out-of-pocket spending and is characterized by a limited insurance system [37].

In 1985, the previously government-subsidized healthcare system was changed to a fee-based cash-and-carry system, truncating the ability of the general population to access formal institutions of care [37]. Pharmacies and chemical shops emerged as key components of health-seeking infrastructure, often taking on a role of community healthcare by disseminating medications and medical advice, with strong potential to fill unmet needs of healthcare institutions [38, 39]. Pharmacies are licensed to sell a variety of medications, some of which require prescriptions or can be distributed at the discretion of a pharmacist [40]. Over-the-counter medicine sellers, widely referred to as chemical shops, are licensed for the distribution of nonprescription drugs [41, 42]. Existing studies point to pharmacies and over-the-counter medicine shops as disproportionate drivers of AMR through excessively lax distribution of medicines as sites of unrestricted, ill-informed antibiotic over-dispensation [43–47]. Distributing antibiotics at high rates can increase the likelihood of their misuse via imprudent consumption for abiotic infections, low-strength dosages, or a consumer failure to adhere to entire treatment courses [48], heightening the risk of evolving drug-resistant disease outbreaks [7, 49].

Prior literature has addressed pharmacies and chemical shops as entities to be studied, with publications written *about* vendors rather than *with* their input, thus failing to highlight the complex experiences of those working within community health contexts (as in most studies addressed in reviews [20, 50]). Recognizing the rich potential of community pharmacy to mitigate outbreaks of preventable and treatable diseases, this study sought to understand the obstacles and barriers to community health provision faced by medicine distributors themselves.

## Methods

### Study setting and context

Data was collected in November and December 2021 throughout 17 neighborhoods in the Greater Accra Region.

As the capital of Ghana, the coastal city of Accra serves as a cosmopolitan node in the network of human and capital flows throughout West Africa. Medicine vendors in Accra are reported to be the primary source of medicine dispensation in Ghana, and in West Africa at large [51, 52]. The city is densely populated and growing each year, a result of regional migration and rapid urbanization [53]. Many of Ghana’s hospitals are located in Accra, and within the city these hospitals are predominantly concentrated in wealthier neighborhoods, a result of colonial segregation and its persistent ramifications in Ghana’s development [26]. Hospitals are reported to be overburdened and crowded, and users describe unexpected fees throughout the care process [54]. The rate of 2.65 doctors, nurses, and midwives per 1,000 members of the population [55] is less than that of the WHO recommendations of 4.45 per 1,000 [56]. Partially as a result, a network of at least 4,198 registered pharmacies and 20,326 licensed over-the-counter medicine sellers (OTCMS) have emerged throughout Ghana [57].

### Survey creation

We first undertook a comprehensive review of the literature, in combination with a series of discussions with stakeholders. Our findings from the review and discussions informed the design of a survey to understand the barriers and facilitators to dispensation of medicines. The survey was piloted at five field sites and revised based on the feedback received. The survey assessed shopkeeper demographics, sales traffic, prices, available brands of antimicrobials, common symptoms observed, and characteristics of the surrounding neighborhood (Appendix S1 File). The present paper relies primarily on responses to qualitative and open-ended questions, with a particular focus on the final question of the survey: “What is the biggest challenge you face as a medicine seller?”

### Definition of study sample

Data was collected in community pharmacies and medicine shops based on a number of criteria, namely the sale of antimalarial or antibiotic medications and the presence of an employee able to participate in the survey. We intentionally curated a diverse set of neighborhoods, in various regions of the city and with a variety of distances from hospitals. Any individual shops we came across in these neighborhoods were invited to participate. Our sampling did not intend to saturate any given neighborhood, but Nima and Maamobi contributed a proportionally higher number of shops. These neighborhoods have been described as low-income and disorganized, so we felt it valuable to oversample these neighborhoods which would otherwise be neglected from the canon of research, to include as much data for the analysis as possible. Shopkeepers were invited to participate by explaining the purpose of the project, providing the informed consent paperwork, and offering a printed copy of the survey to aid in deciding whether to participate.

### Inclusion and exclusion criteria

Any employee over the age of 18 who sold antibiotic and/or antimalarial medications in an herbal shop, chemical shop, or pharmacy in the Greater Accra region was eligible for participation in English or Twi.

89 shops were visited, and 94% (n=83) agreed to take part in the study. Of the respondents who offered to participate, 80 met the inclusion criteria and completed the survey in full. Forty-five of the participating field sites were pharmacies, 32 were chemical shops, and three were herbal shops.

### Ethical considerations

The study was approved by the New York University Abu Dhabi Institutional Review Board (HRPP-2021-146). A detailed consent form was distributed to participants after introducing the project verbally. Shopkeepers were invited to decline to answer any questions and withdraw at any time. Some surveys were administered on paper, others on a laptop using Qualtrics. The Qualtrics questionnaire automatically skipped questions about antibiotics for chemical shops to investigate only the over-the-counter medicines they were licensed to sell. No identifying information of shops or of participants were recorded, and location data was noted only at the neighborhood level. For confidentiality purposes, surveys were conducted in a location directed by the participant. Surveys were administered verbally and simultaneously typed on a laptop or written on printed copies and later transcribed. Participants were compensated 1.00 Ghanaian cedi per minute that the survey required (USD10.00 per hour).

### Conceptual framework

Shopkeepers’ roles in providing community care, through dispensation of medicine, were contextualized via the socioecological model of public health. This model, first delineated by McLeroy and colleagues in 1988, categorizes health-influencing components at the individual, interpersonal, organizational, community, and public policy levels [58]. The model posits that examining these factors and how they interact and overlap, can be essential for developing interventions and strategies to improve health and wellbeing. Individual characteristics could include beliefs, attitudes, and behaviors influenced by personal background. Interpersonal factors on the other hand are defined as the relationship between main stakeholders (in this case distributors and customer-patients). The community level includes neighborhood conditions, focusing on the relationships among organizations and groups within it. Organizational characteristics include the structuring of institutions within the healthcare system, and public policy encompasses the broader political and economic changes affecting community pharmacy. We examine the various elements of the model to identify barriers, and determine the contributions of these factors to the challenges.

Responses to the question “What is the biggest challenge you face as a medicine seller?” were compiled for all 80 participants. Responses were coded by one reviewer using a qualitative content analysis method, and 20% were independently coded by a second reviewer. The extent of agreement between coders was k= 0.95. Each response category was then correlated to one or more of the five tiers of the socioecological model of public health, with the number of responses in each category and each tier of the socioecological model tabulated. All responses which alluded to challenges and barriers were compiled in a list. A snowball method was used to select additional qualitative survey questions for analysis; if the response given by participants describing their largest challenge was related to a separate question, all relevant responses to the additional question(s) were considered for inclusion in the analysis. We also present demographic characteristics of respondents, which were analyzed using Stata.

## Results

Respondents included 17 pharmacists and 42 Medicine Counter Assistants (MCAs), and the average length of training undertaken prior to employment had been 19.6 months (SD=21.7). The majority of respondents were under 34 years of age and had been working in the field site for five years or fewer. 72.4% of respondents were female. For complete demographic information about the n=80 respondents see Table 1.

**Table 1.**
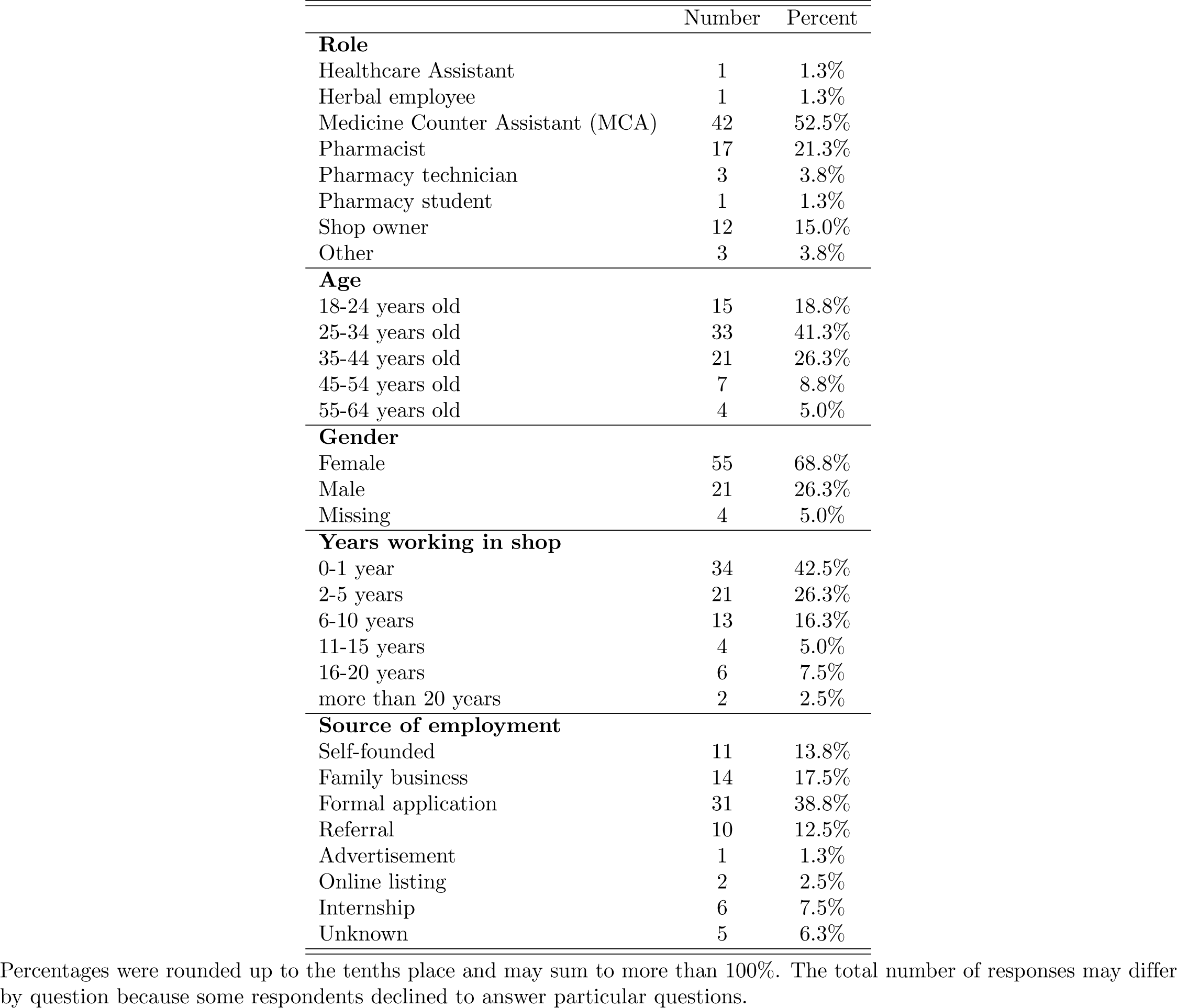
Demographic characteristics of respondents.

Table 2 presents occupational information about shopkeepers’ work. Participating respondents had worked in their shops for an average of 5.11 years. Shopkeepers reported working an average of nearly ten hours per shift, six days per week, suggesting a 60-hour workweek. Each shop employed between 1 and 25 employees, with an average of 4.19.

**Table 2.** Shopkeeper work experience information

| Type |  | Mean | SD | Minimum | Maximum | N |
| --- | --- | --- | --- | --- | --- | --- |
| **Chemical/herbal shop** |  |  |  |  |  |  |
|  | Number of months of training | 11.7 | 12.4 | 0.0 | 48.00 | 29 |
|  | Number of years working in this shop | 6.5 | 8.3 | 0.0 | 37.0 | 35 |
|  | Year of starting to work in this location | 2014 | 8.30 | 1984 | 2021 | 35 |
|  | Number of days worked each week | 6.4 | 0.6 | 5 | 7 | 35 |
|  | Number of hours worked in each shift | 12.0 | 3.9 | 1 | 24 | 32 |
|  | Number of hours shop is open each day | 14.3 | 2.9 | 9 | 24 | 35 |
| **Pharmacy** |  |  |  |  |  |  |
|  | Number of months of training | 25.35 | 25.15 | 6.00 | 72.00 | 40 |
|  | Number of years working in this shop | 4.07 | 5.29 | 0.00 | 19.00 | 45 |
|  | Year of starting to work in this location | 2016 | 5.29 | 2002 | 2021 | 45 |
|  | Number of days worked each week | 5.98 | 0.76 | 5.00 | 7.00 | 44 |
|  | Number of hours worked in each shift | 8.23 | 1.37 | 6.00 | 12.00 | 41 |
|  | Number of hours shop is open each day | 15.49 | 3.79 | 10.50 | 24.00 | 43 |
| **Total** |  |  |  |  |  |  |
|  | Number of months of training | 19.61 | 21.72 | 0.00 | 72.00 | 69 |
|  | Number of years working in this shop | 5.11 | 6.81 | 0.00 | 37.00 | 80 |
|  | Year of starting to work in this location | 2015 | 6.81 | 1984 | 2021 | 80 |
|  | Number of days worked each week | 6.18 | 0.71 | 5.00 | 7.00 | 79 |
|  | Number of hours worked in each shift | 9.89 | 3.34 | 1.00 | 24.00 | 73 |
|  | Number of hours shop is open each day | 14.97 | 3.45 | 9.00 | 24.00 | 78 |

The total number of responses may differ by question because some respondents declined to answer particular questions.

10% of shopkeepers reported that they do not perceive any substantial challenges (n=8). An average of 1.68 and maximum of 4 distinct challenges were reported by each respondent. Shopkeepers reported a total of 128 obstacles they faced in their role as providers of medicine, and these included individual, interpersonal, community, organizational, and public policy level factors. The categories developed for the socioecological model are depicted in Fig 1.

**Fig 1.**
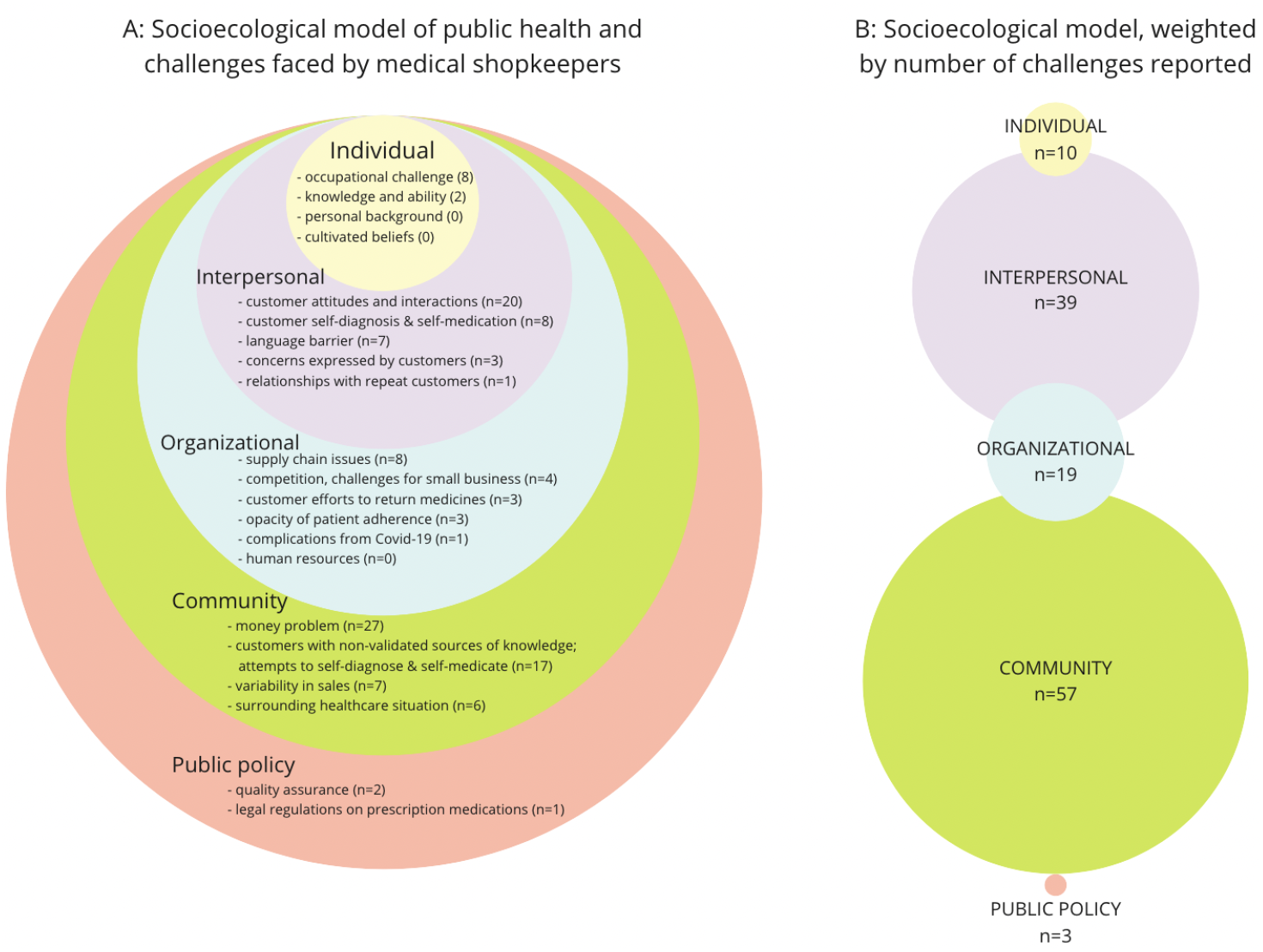
Challenges reported. **Panel A:** Participant responses as categorized into the socioecological model of public health. Note that 42 shopkeepers mentioned more than one challenge. **Panel B:** Categories in the socioecological model of public health, weighted by the number of challenges reported. The diameter of each tier is proportional to the number of responses in that category.

The obstacles reported as the primary challenges shopkeepers faced in their work distributing medicines were tabulated by category and represented by the diameter of each circle weighted by the number of responses, as depicted in Fig 1.

### Individual

Shopkeepers referenced a range of distinct personal backgrounds, education routes, beliefs about community health, work characteristics, and sources for acquiring further knowledge. Not all categories were necessarily identified as challenges, in contrast with prior studies. Categories which were theorized in prior research but unreported by participants in the present study appear in Fig 1 as (n=0). Respondents’ degree of enthusiasm for their occupation and its challenges varied.

Among the employment-related challenges cited by respondents were their insufficient income (specifically listed as the biggest challenge by one), a language barrier with customers (n=7), boredom (n=1), and not “having enough time to counsel each patient” (n=1).

Most shops sold a variety of artemisinin-based combination therapy (ACT) antimalarials, and pharmacies in particular tended to sell a variety of both imported and local brands of antibiotics. Shopkeepers reported an absence of centralized guidance about which of these brands were in fact most effective, though some reported feedback from returning customers. Many shopkeepers had thus developed individual opinions about each brand through feedback about patients’ experiences; some shopkeepers voiced a stronger belief in the efficacy of imported or originator brands than local generic brands, with one saying “you can never compare generics to innovator, [imported originator brands being] more efficacious.” These opinions were based on limited patient feedback, rather than centralized system-wide data, potentially impeding prudent distribution.

Shopkeepers’ degree of concern with antimicrobial resistance varied widely, but many noted a strong belief that antimicrobials had to be sold in full and completed as entire treatment courses, guarded for only the cases they deemed necessary. However, they reported a general uncertainty about the issue and a lack of clarity on the state of antimicrobial resistance in Ghana. To overcome this lack of centralized guidance, one pharmacist reported independently reading scientific articles to remain informed about current best practices, and others reported doing research on the internet. Informants explained that AMR was not specifically considered before distributing medications, and some seemed to worry about the rapid emergence of drug resistance rendering their products obsolete, without centralized guidance on which drugs should be still used.

### Interpersonal

Interpersonal challenges included the attitudes and concerns of customers and their efforts to self-medicate.

When asked about the greatest challenge they faced, many shopkeepers complained of customers who were rude, made insults, or initiated arguments. Some respondents referenced their need for a great deal of patience, and the proverb “the customer is always right” was mentioned repeatedly, sometimes with chagrin.

The heightened emotions associated with illness were reported to exacerbate shopkeepers’ challenges, as articulated by one: “People walk in with anger. When sick, people become frustrated. They ask for a certain drug… Some may insult or say mean things… I try to advise, but they don’t want to be told what he/she should consume.” Many shopkeepers reported customers resisting their inquiries into symptoms or dismissing their advice. Regarding their stewardship of antibiotics, one reported “Some patients don’t understand antibiotics for instance. [Customers say] ‘ I am buying, [I] don’t understand why you don’t want to sell it to me.’ Sometimes it turns into an argument.” Shopkeepers reported being caught between meeting patients’ demands and retaining commercial viability, while also denying sales to safeguard medications they deem inappropriate. Many cited difficulties along the lines of “trying to get them to understand why they don’t need an antibiotic for a very trivial issue” while occupying a role as health educators in a status position that is typically seen as lower than doctors.

The practice of self-diagnosis was reported to be widespread, whereby customers subverted the process of going to hospitals or clinics and instead identified an illness themselves based on the symptoms they perceived relevant. Efforts to self-medicate with over-the-counter medications often brought customers to pharmacies or chemical shops. When customers believed they needed prescription medications such as antibiotics, shopkeepers reported difficulties: “customers tell ‘ I want this’ but this medicine will not solve their problem. Sometimes they will not understand why, but I say no [to their request] if I know it won’t help them.” Shopkeepers alluded to their conditional allowance of customer self-medication, perhaps offering medications without prescription if they deemed the chosen treatment course useful to the customer. To protect subjects’ confidentiality, we did not ask explicitly about illegal antibiotic distribution. Thus, inferences were made and extrapolated from available responses such as over-the-counter antimalarials.

When shopkeepers tried to explain to customers that a requested medication was not prudent or instead offered an available brand with the same active ingredient, some customers were reportedly staunch in preferring one particular variety.

Some shopkeepers mentioned the potential for drug abuse; one believed that it occurs “especially [for] antibiotic and analgesic” medications, and another explained that they responded to such situations by dodging the customer’s request: “If I think they will abuse it, I tell them it is out of stock.” Though it was unclear what was explicitly interpreted as “abuse”, the context of antimicrobials suggests that abuse could involve either taking an excessive dosage at once, or using medicines for effects other than their prescribed purpose. Future work should elucidate the contextual meaning of “abuse” and its different implications for successful interventions.

Respondents reported that challenges arose when family members who were ill would send children to purchase medication. With conditions of poverty and an inability to miss work, children often served as interlocuters to retrieve medicines. In response, shopkeepers reported that they would send the child away, require a written note from the parent to clarify the correct medication, or call the parent to confirm an over-the-counter medication in case the child misspoke. Shopkeepers reported that dealing with children made providing quality care much more difficult.

80.0% of shopkeepers (n=64) believed that many of their customers lived in the neighborhood surrounding their shop. Some shopkeepers proclaimed a lack of challenges, attributing this to their integration in the surrounding community; several echoed the quote that they “have not faced much challenge due to familiarity.” These shopkeepers may have also experienced the challenges listed by other respondents, but did not perceive them as major challenges worth mentioning. Occupational challenges may have been framed differently in the eyes of long-serving shopkeepers, who considered such obstacles to be ordinary workplace conditions taken for granted.

70% (n=56) of shopkeepers reported that most customers returned to their shop multiple times, while only 7.5% (n=6) reported that most customers visit only one time and do not return. Upon return visits to a given enterprise, some customers reportedly expressed their concerns to shopkeepers. The single most common challenge described by shopkeepers was that of money; 53 respondents (66.3%) alluded to their customers’ payment and the cost of medications as an obstacle. Prices varied by shop, with the overall selection of antimalarials ranging from $0.50 for local varieties to $7.00 for an imported brand. These costs were reported to strain financial resources and be prohibitively high for some customers.

Most shops explicitly specified that they required payment immediately at the time of sale; eight reported as their biggest challenge “customers trying to buy on credit.” The practice of credit occurred only occasionally, dependent on the individual shopkeeper. Some reported that they would offer medicines on credit and trust that a person would return to pay later. This was more common for those who reported being familiar acquaintances with repeat customers. One reported that for patients who were “struggling and can’t pay, I give and pay the rest out of my own funding.”

Some shops reported a lack of diagnostic testing altogether, and others explained that rapid diagnostic testing for malaria was only available for those who “had extra money” for the out-of-pocket fee of diagnostic testing. They suggested that most customers did not opt for the extra cost and instead pursued treatment for presumptive malaria based on symptoms. Some shops reportedly required a rapid diagnostic test result before they distributed antimalarials. One reported that they previously operated on presumptive positive treatment based only on symptoms, but after beginning to offer rapid diagnostic testing, the number of antimalarial medications they sold plummeted. This suggested that they were previously over-distributing antimalarials for cases that were not actually malaria.

Side effects were not mentioned often in customer complaints, but one shopkeeper reported “[I] don’t do malaria tests because injections are not liked,” indicating that customers’ disdain for blood sampling led to the blind treatment of malaria based on symptoms rather than diagnostic testing. Regarding customer concerns about side effects of medications, another respondent explained “only one out of ten of them has complained,” and another said “For other medicines, people complain about side effects, not really [for] antibiotics.”

The availability of dosage strengths and treatment options were described as challenges for some shopkeepers. Medications that required frequent dosages (such as every four hours, or tablets with small dosages that required taking up to four pills at each interval) were reportedly disliked by customers. Some customers would decline to purchase these medications in the first place, and shopkeepers expressed concern that patients who did purchase these typically lower-cost medicines likely would not adhere to their regimens.

Shopkeepers reported that many customers established preferences for particular medications based on prior experiences, advertisements, or reports from acquaintances about which medicines effectively cured their ailments. Many shopkeepers reported with consternation that television and billboard advertising determined which brands customers preferred, with one shopkeeper stating that their biggest challenges was “trying to convince a patient to buy another type of drug aside from what he or she thinks is good or what a layperson or TV/radio advert[advertisement] has explained.”

### Organizational

Regulations and structures governing the provision of healthcare may have generated challenges in human resources, patient monitoring, and supply chain management. Economic competition, responses to the Covid-19 pandemic, and the proximity of shops to other medical institutions were reported to have reduced sales for many shops.

Within each shop, particularly in larger enterprises, some respondents described a hierarchy of employees’ roles shaping the provision of medications. As reported by participants, the pharmaceutical sector in Ghana is regulated by a tiered system of employee titles and prescription requirements. Under pharmaceutical regulations, licensed pharmacists can distribute restricted classes of medicines (including some antibiotics) at their own discretion when patients do not have prescriptions. On the other hand, Medicine Counter Assistants, who complete six months of training, do not have this level of agency and are only supposed to recommend over-the-counter medicines or distribute based on existing prescriptions [40, 59]. MCAs may defer to pharmacists when navigating customer demands for higher-tier medications, but the pharmacist may be present only for part of the day, reportedly often the evening shift. Facing an unmet need of licensed pharmacists, other employees may take on pharmacist-specific tasks despite having limited training. Some MCAs explained that they told patients to wait and return to the shop later in the day when seeking medications that could be prescribed by a licensed pharmacist, but it was implied that they may sometimes distribute antibiotics at their own discretion. The disparity between licensed roles and actual practice was thus mentioned as a challenge by some respondents.

Shopkeepers mentioned the absence of a system for following up with patients and monitoring their compliance with treatment courses, particularly for antibiotics. Some respondents believed that patients likely did adhere to their treatment because “it is expensive, no one wants to waste money,” while others were less certain: “might or might not; we are not able to track over time.” The majority echoed this sentiment, and expected that customers would *not* complete their treatment courses, with only 41% of shopkeepers believing their customers “probably” or “definitely” did adhere to their entire antimicrobial treatment courses. Adherence monitoring was described as being undertaken at an individual level. When asked “Do you think customers actually finish the entire treatment course of antibiotics after they leave the shop?” one responded “Definitely yes, because I talk to them. I tell them when you don’t finish up with the course, your treatment will not wash off [the infection].” Another reported that they took phone numbers and routinely called patients to verbally ensure that they finished their antibiotic course.

Some shopkeepers reported the competition of “too many pharmacies” as their greatest challenge. Price comparisons sought by customers were reported to favor larger chain enterprises rather than smaller shops. Many franchised retail pharmacies also functioned as wholesale suppliers, even manufacturing their own ACT antimalarials through an affiliated factory. A respondent explained that wholesale suppliers calculate costs based on the number of medicines purchased, so shops “taking just five packs of Lufart” (a locally-manufactured brand of antimalarial), for example, would pay more per unit than chain enterprises purchasing large quantities.

45 shops reported no delay between running out of a medication and resupplying, and a further 30 reported that medicines could be restocked within a matter of days. Still, shopkeepers reported some challenges with the supply of certain brands patients preferred and with quality assurance. For instance, several respondents complained that medicines would arrive at or close to their expiry date, and with low sales traffic would expire before they could be sold. Reports of shopkeepers’ greatest challenges included “low sales” and the “expiry of drugs; when drugs arrive they are almost expired and don’t move fast.”

Many shopkeepers reported that the Covid-19 pandemic reduced their sales and customer traffic. Respondents theorized that, fearing both a potentially infectious environment and the social and economic repercussions of a positive Covid diagnosis, many would-be customers avoided sites they associated with sickness. Asked of broader annual fluctuations in customers, a pharmacy shopkeeper replied “Sales are low.

Malaria symptoms are just like Covid symptoms. If [they] have malaria, just similar to Covid— cough, sneeze, feverish, just the symptoms of Covid. It’s difficult because of this Covid. It interrupted a lot of pharmacies because [would-be customers are] afraid to come for drugs.”

### Community

Broader challenges faced in the community were reported to intersect with a lack of awareness about medications and a lack of proximity or access to other healthcare services.

A lack of customer/patient awareness of antimicrobial stewardship was described by some shopkeepers, which they often attributed to low education or literacy. Even when customers had awareness of proper use, they may have lacked access to other healthcare services because of prohibitive costs or geographic distance from relevant medicine.

Shops’ proximity and patients’ differing access to hospitals and clinics were reported to heavily influence customer traffic and behaviors. A pharmacy within a hospital reported that patients often chose to take their prescription notes and buy medicines elsewhere for lower prices. Many shops were observed to cluster around hospitals, and a price-comparison practice was reported, in which customers started at the pharmacy closest to the hospital, inquired about prices, and moved down the line to adjacent pharmacies until they found the lowest price. Customers’ purchasing decisions thus appeared to be made based on price above any other factors.

One shopkeeper reported as their primary challenge the insufficient “availability and accessibility of diagnostic tests and diagnostic facilities for customers or patients.” Conversely, some shops reported that they did *not* offer diagnostic testing for malaria because they could be accessed at the hospital nearby. Shops in neighborhoods surrounding hospitals reportedly relied more heavily on prescriptions than on symptom-based requests from walk-in patients. Proximity to a hospital was reported to affect the reliance on prescriptions and customers’ ability to afford medicines. Shops near hospitals faced less difficulty with customer finances and relied on prescriptions more, because their patients were more likely to have undergone diagnostic testing.

The ability to access hospitals was reported to sometimes predict customers’ medication purchases. Three shopkeepers asserted that visiting a hospital or doctor was an established indicator of affluence. When asked “What would you do if a person doesn’t have enough money for the antimalarial/antibiotic they intended to buy?” a shopkeeper working in close proximity to a hospital responded “Usually they have the money because they’ve been to the hospital. Pharmacy is the first point of contact in the community. If you can [afford to] take a cab to a hospital or clinic, you can afford it [the medication].” This suggests that medicine shop customers without prescriptions were seen as less affluent customers, lacking the financial resources to confer the ability to access doctors or purchase medications easily. Articulating the disparity in access to medicine, one respondent categorized the residents of their surrounding neighborhood as “elite” and said that their clients preferred calling a doctor rather than relying directly on a pharmacist. A higher baseline of wealth reportedly enabled access to doctors in formal institutions of care, while lower-income customers had only sporadic access to care and may have been seen as less reliable customers.

Medicine shops reported encountering fluctuations in sales based on broader economic and political changes. The high rate of informal employment in Accra was reported to create financial variability within customers, potentially undermining the stability of medicine shops and their ability to regularly procure medications. Medication purchasing appeared to be characterized by economic instability: when asked to rank the income level of residents of the surrounding neighborhood (low, medium, or high), answers varied within neighborhoods and one shopkeeper answered “sometimes low, sometimes high.”

### Public Policy

Reported challenges with potential policy implications included diagnostic capacity, prices, supply management, and drug quality assurance. However, direct government involvement was not proposed or mentioned by participants, many of whom echoed a general disillusionment with policymaking and an absence of expecting government support or action on the concerns they face.

## Discussion

Employees in pharmacies and chemical shops have created an informal system of healthcare at the neighborhood level, appearing to perform a vital role in community health as providers of medical care filling in gaps left by larger institutions. In their role, however, they found a number of obstacles that truncate their ability to fully meet the needs of patients.

The background and beliefs of a medicine vendor may have occasionally influenced whether and how medicines were distributed because the relatively unregulated medical system could leave space for individual perceptions to dictate unorthodox antimicrobial use. On the whole, however, the surveyed shopkeepers reported efforts to be judicious about the antibiotics they gave. Their overall desire to safeguard medications appeared to be an improvement from the findings of previous literature [44, 60].

Despite their intentions, a number of factors still complicated shopkeepers’ abilities to practice stewardship. These challenges were situated throughout the socioecological model of health, but were more concentrated at the community and organizational levels. This was in contrast to the aforementioned prior literature, which pointed to individual-level agents as drivers of antimicrobial misuse. This suggests a need for realignment of focus in the field.

After mapping shopkeepers’ reported challenges within the socioecological model of public health, three themes emerged as the primary challenges affecting stakeholders: (I) serving as health educators and gatekeepers of medicines while operating within a fragmented system of healthcare; (II) the economic ecosystem with cost as the major limiting factor of health-seeking behaviors; and (III) direct challenges in the treatment of illnesses.

### (I) Challenges to a role as health educators for the community and gatekeepers of medicines

Though Medicine Counter Assistants and pharmacists were not fully sanctioned as healthcare providers, many reported feeling the need to fill a void created by the difficulties that patients face in trying to visit hospitals. This reaffirmed prior literature citing frequent self-medication through private medical shops in Ghana and beyond [37, 41, 61–64].

Shopkeepers’ individual challenges, such as insufficient income, may play into their enthusiasm for the job and the capacity of care they are able to provide. With over 50% of shopkeepers employed less than two years in their present workplace, there appear to be high rates of employee turnover. Individual-level factors may affect the likelihood of long-term career attrition. Especially influential for community health, long-serving shopkeepers may become embedded in the community as trusted health providers, recognizing patterns to establish best practices in care. Conversely, they may retain outdated conceptions of medical practices. In the context of AMR, the rapidly evolving situation requires frequently reevaluating which medicines are to be avoided; if resistance develops like it has for penicillin and chloroquine, it is unclear whether prescribers are given updated information or continue distributing the medications originally encouraged decades prior.

#### Decision-making on antibiotics: to medicate or not to medicate?

A lack of awareness of AMR and a high level of comfort with antibiotics appears to drive many customers to specifically request antibiotics even without the diagnosis of a bacterial infection. Shopkeepers described such requests as challenges when contradictory to their judgment of the utility of antibiotics. Shopkeeper responses suggested that complicated balancing acts weighing risks and benefits were at play.

Even when a bacterial or malarial infection was not confirmed via diagnostic testing, antimicrobials were still sometimes judged to be the most economical solution. The calculus underlying such decisions based on community health challenges was initially spurred by the individual patient, then sometimes altered with the advice given by shopkeepers [65, 66]. Even if the long-term risk of AMR is known, the risks and implications of *not* taking antibiotics—and thus leaving a disease untreated—could be enough to drive their unprescribed use [67].

#### Public policy implications

Overall, it appears that antimicrobials are now being more carefully guarded than they were during prior studies [41, 47, 60, 68–70]. Existing literature claimed that vendors distributed antimicrobials regardless of whatever laws were in place, asserting that this was a widely known expectation among the general public [44, 60]. Indeed, many customers seemed to expect easy access to antibiotics; as reported by one shopkeeper, “If they want [an] antibiotic and you’re asking questions, it’s like ‘ hey madam, why you asking plenty questions,’ like that, ‘ I just want antibiotic, why don’t give it to me and go?’ “ This could be indicative of customers’ prior experiences of abundance contradicting the present-day legally regulated stewardship to which most respondents appear to indicate strict adherence. It is worth noting, however, that survey questions were designed not to require divulging any noncompliance with legal restrictions, in order to mitigate the risk of participating in this research. Patients’ efforts to self-medicate in the existing logistical system often compel shopkeepers to act as fill-in healthcare providers. However, legal regulations sometimes truncate their ability to distribute as intended. This can present ethical and legal dilemmas if shopkeepers seek to distribute prescription medications at their own discretion.

Policy changes in Ghana have focused on restricting the availability of antibiotics through prescription laws, but our study suggests that the most significant challenges driving AMR are not fully addressed by these policies. The major challenges neglected by public policy appear to include a lack of diagnostic testing and the financial inaccessibility of full-course medicines.

Shopkeepers who recently adopted rapid diagnostic testing reported selling antimalarials less often than they previously had when treatment routes were based only on symptoms. This reaffirms prior studies detecting high rates of overdistributing antimalarials for non-malaria cases [71–73]. The more prudent use of antimalarials after the introduction of rapid testing encourages upscaling access to diagnostic testing for malaria in order to prevent the overuse of antimalarials and accelerated resistance.

### (II) bEconomic ecosystem and cost as the most significant barrier

In their efforts to maintain competitive prices, shopkeepers cited challenges in supply, procurement capacity, prices, and factors disrupting the larger economic ecosystem, such as competition and the Covid-19 pandemic.

#### Prices

The cost of treatment was reportedly the primary concern voiced by customers, many of whom found medications to be prohibitively expensive. Cost was reported to supersede other factors and shape how customers interacted with distributors on an interpersonal level and with the healthcare system more broadly.

The shops which refused to sell partial, subtherapeutic treatment courses may have lost customers in their efforts to practice antimicrobial stewardship, because customers may have left and sought one of the noncompliant shops which would sell piecemeal partial treatment courses of antibiotics. Thus, a concerted effort must integrate *all* shopkeepers and strengthen their capacities to navigate the stewardship of antimicrobials without jeopardizing the financial security of individual gatekeeping shops.

Capacity-building for shopkeepers may aid in heightening their status in their communities to leverage a role as gatekeepers without financially jeopardizing their enterprises. While physicians are typically held in high regard at the top of the healthcare provider hierarchy in Ghana, shopkeepers may not be perceived as experts or may be disregarded if they try to act as stewards of particular medications.

#### Complications from the Covid-19 pandemic

The broader trend of economic downturn in Ghana due to Covid-19 [74] intersected with medicine dispensation in these sites in ways that were not expected. Because of the inability of the general public to access healthcare or additional efforts to avoid overcrowded hospitals during the outbreak, there could have emerged an increased reliance on pharmacies and non-institutional settings for self-medicating ailments [75]. Post-pandemic health-seeking behaviors may have shifted away from institutions of formal health care and further into self-medication practices (as seen in [15, 76–78]).

One could expect that a disease outbreak would spur higher need for medications or protective supplies such as face masks or disinfectant. Undiagnosed cases may have relied on pharmacies or chemical shops for symptomatic treatment using over-the-counter medications. Unexpectedly, however, many shopkeepers in the present study reported that their sales declined as the outbreak spread. Shops found their role shrinking due to clients’ fears of infectious spaces and their anticipation of repercussions following a Covid-positive diagnosis. As a shopkeeper reported, “They will not come for a drug thinking they might be told they have Covid.” As seen in such responses, a positive test result would lead to stigma and missed days of work, creating logistical, social, and financial consequences such that many people may have sought to avoid medicine shops altogether to avoid the possibility of being diagnosed.

Many respondents pointed to a particular reduction in malaria medication sales, believing that customers did not want to be told that they had Covid-19 upon visiting a pharmacy if asked to identify their symptoms, which shopkeepers described as similar to malaria.

The Covid-19 pandemic highlighted the vulnerability of healthcare systems, and this study illustrates the multifaceted challenges faced by workers who are not as visible as the doctors and nurses in formal institutions of healthcare. Particularly in contexts with high rates of unmonitored medicine use, strengthening the existing infrastructure appears critical to mitigating future outbreaks of antimicrobial-resistant pathogens.

### (III) Treatment-based challenges

Shopkeepers consistently mentioned two main organizational-level challenges limiting the effectiveness of the treatment that they provide: patient nonadherence to medications, and the supply of efficacious medicines. Although these were not often voiced as the single largest challenge faced, these obstacles could have particularly profound impacts driving the risk of antimicrobial resistance.

#### Adherence to treatment courses

Firstly, the monitoring of adherence to antimicrobial treatment courses is done at an individual level, rather than systematically. Shopkeeper challenges include a low ability to follow up on patient adherence, and interventions to improve the continuity of care at the community level could be highly useful. Many shopkeepers reported doubts about their customers’ knowledge of the risks of antimicrobial misuse and subsequent completion of their treatment courses. Shopkeepers reported that their private medical enterprises were challenged by customers’ inabilities to afford diagnostic testing, formal medical guidance from physicians, or the gold-standard antimicrobials of proven efficacy. These obstacles echo the findings of prior studies in low- and middle-income countries [14, 48, 66, 79–81].

#### Efficacy and supply of medicines

Supply chains were reported to be relatively reliable, in optimistic contrast to prior literature [48, 82]. Shopkeepers voiced a recurrent preference for imported ‘ originator’ brands of medication, which many believed to be more effective than locally-manufactured varieties. Imported medicines were, however, typically more expensive and could sometimes be more difficult to procure. Lower-cost local brands may have been more accessible and could match their efficacy, though there was limited infrastructure in place to guarantee their quality [83–87]. Some shopkeepers may have been interested in relevant government bodies supporting improved procurement and quality monitoring, in order to offer effective medications at prices their customers could afford.

### Limitations

Social desirability bias and the potential legal ramifications of responses may have led participants to give idealized responses to be compliant with expected standards.

Additionally, responses may have been affected by the positionality of the researchers. The identity of each researcher, one a white U.S. American and the other from Northern Ghana, may have skewed the level of trust which was cultivated and participants’ openness to answering. The participation rate was almost identical for each field researcher, but there may have been differences in communication and distinct types of desirability bias. While this study sought to be as self-reflexive as possible, social dynamics, problematic histories of white researchers, and axes of privilege may have shaped responses and our subsequent interpretations. This study recognizes that responses may have been biased by these dynamics, and results should be interpreted in that lens. Any extrapolation of these findings is limited by its status as research operating at a distance from the experiences of those directly affected. These findings are offered as only one possible interpretation of complex dynamics, cultivated through a research setting rather than firsthand lived experiences.

The subjective nature of some survey questions may have skewed responses, particularly if they morphed slightly or were interpreted differently by respondents. The questionnaire was designed to build upon and corroborate answers to interrelated questions, but the potential for misinterpretation and mistranscription remains. Some responses may have been more anecdotal than representative if recent experiences determined what was most salient to shopkeepers and prone to be reported. Shopkeepers’ beliefs about consumers’ behaviors *after* purchasing medications and leaving their shop may be inaccurate.

Research in this context builds on an extensive history of problematic practices, and this presence as a white researcher may perpetuate colonial hierarchies. No analysis would be complete without contextualizing the lack of healthcare accessibility in Ghana today at its historical, colonially-driven source, and this study should be interpreted in this larger context. If shopkeepers are ill-informed or face challenges because their healthcare system is fragmented, it may not necessarily be the fault of individual shopkeepers or the healthcare system itself, but rather the colonially broken foundations and conditions imposed by international systems [37].

### Future directions

Replicating the present survey in a variety of contexts can improve broader global understanding of antimicrobial distribution. Within Accra, surveying more chemical shops and pharmacies across a wider range of neighborhoods, supplemented with longer-term observational studies, could help validate the quantitative findings of this limited sample size. Laboratory testing to determine the efficacy and quality of the ingredients in available medications would offer essential insights into how supply chains and medicines are performing.

Pinpointing the context-specific drivers of antimicrobial misuse can help allocate resources to target specific practices shown to accelerate drug resistance. Antimicrobials are critically important life-saving medications, but because of heavy reliance and the lack of new innovation, medicine has been losing the race to microbes evolving resistance. To promote stewardship, it is essential to fully understand the challenges plaguing grassroots distributors of antimicrobial medications. The capacity-building of community medicine distributors has been relatively neglected and should be studied further. For contexts struggling with neglected tropical diseases that can be treated with straightforward regimens, existing community health distributors may be valuable stakeholders with extensive potential to improve community health. Expanding safe diagnostic capacities to these informal community health providers should be investigated for viability as a measure to alleviate the burden on crowded hospitals, improve treatment outcomes of the general population, and better inform the prudent use of antimicrobial medications.

With high percentages of customers expected to live in the neighborhood surrounding each shop, shopkeepers may provide a substantiated glimpse into the public health situation at the community level. Broader challenges faced in the surrounding community, such as limited healthcare access and prohibitive costs of formal healthcare institutions, directly shape the experiences of these medicine enterprises (as similarly discussed by Buor and colleagues [88]. Shopkeepers’ reports suggested that they are intrinsically embedded in the communities they serve on a daily basis, thus establishing rapport, hearing concerns, and learning of treatment outcomes. Confirmatory studies should examine long-serving medical shopkeepers’ knowledge of recurring trends in symptoms and treatment patterns to determine how and whether they may wish to be mobilized as key informants of epidemiological surveillance (as in the ABACUS project by Wertheim and colleagues [89]).

#### Recommendations

1. **Free rapid diagnostic testing:** Providing confirmatory diagnostic testing without additional fees appears extremely important for ensuring the judicious use of antimicrobials. Increasing the availability of tests and subsidizing fees for their users could enable more informed treatment of malaria based on diagnoses rather than symptom-based presumptive treatment. This may require more laboratories or subsidies for rapid in-store diagnostic tests.
2. **Quality assurance for customer trust:** Shopkeepers reported that many customers prefer to use only brands that they are familiar with, founded via prior experience or the recommendations of trusted acquaintances. The general public may recall stories of counterfeit and substandard medicines in West African markets [42, 90], driving valid concerns expressed by many customers when offered medications they do not recognize.
3. **Minimum strength for active ingredients:** Monitoring and enforcing the efficacy of medications is critical to ensure that antimicrobials are of sufficient quality and strength to mitigate opportunities for microbial evolution. Enforcing mandatory minimum strengths for medicines could help prevent the use of subtherapeutic doses by making sure all medicines in inventory are sufficiently efficacious.
4. **Include the active ingredient in advertising:** Encouraging the listing of active ingredients in advertisements could help customers recognize the legitimacy of medicines offered, rather than depending on only one particular familiar brand. As a shopkeeper reported, “If you don’t know [this medication] and I give you, it will worry you.” More tightly regulated monitoring may assuage these fears and could help promote local manufacturing.
5. **Shelf stability:** The development of more shelf-stable medications can help mitigate the untimely expiry of medicines, as could a consignment system for manufacturers to replace drugs that have expired. Given that none of the visited shops appeared to have refrigerators for medications, it is possible that their expiry may occur faster than expected. Thus, it should be ensured that medicines are shelf-stable without refrigeration.
6. **More robust reporting pathways:** Based on shopkeepers’ reports, it is apparent that centralized monitoring and a unified information source for antimicrobial resistance should be developed for physicians, pharmacists, and MCAs to communicate when antibiotics do not work or indicate projected drug resistance. Streamlined routes for reporting side effects could also improve customer trust and promote the availability of safe, effective treatments.

## Conclusion

The organization- and community-level tiers of the socioecological model of public health presented the most profound challenges for the health-providers studied in Accra. Based on their reports, antimicrobial misuse practices in Accra appear to be not the result of individual misbehavior or gaps in knowledge, but instead indicate systemic and recurring gaps in the ability to access effective medical treatment.

Prior AMR research in Accra has centered on drug-resistant outbreaks in hospitals, [91], but the ability to attend a hospital may sometimes be predicated on a level of privilege that neglects populations unable to access such institutions, and thus can leave the most devastating outbreaks out of the purview of these studies. Communities without access to formal healthcare appear to seek cheaper independent access to pharmaceuticals rather than consulting physicians in formal institutions [54, 64, 85, 92]. Future research should continue to examine the importance of medicine distributors in the community, believed to be a source of a large percentage of all antimicrobials used.

While AMR is a global issue, its repercussions are likely to fall most heavily on the communities least prepared to mitigate such outbreaks. Significant populations seek care outside of formal healthcare institutions and are thus neglected in estimates of disease prevalence; partnering with the informal healthcare sector could improve the accuracy of AMR reports and the quality of health care by more robustly assessing and monitoring global health [2, 18, 93, 94].

## Data Availability

The data underlying the results presented in the study are available upon request by contacting the corresponding author.

## Supporting information

**S1 File. Survey questionnaire**. The complete survey questionnaire.

## Acknowledgments

Above all, we thank the participants of this research who shared their experiences and generously shared time out of their workdays, going above and beyond to explain the landscape of public health through their eyes.

This fieldwork would not have been possible without Rafia Abdul-Mumin, who conducted 40 fieldsite visits. Her insight and hard work were invaluable, and the success of the study is truly indebted to her.

The fieldwork for this project was funded by the Social Sciences Division at NYU Abu Dhabi, and the publication was funded by a grant from the Effective Altruism Infrastructure Fund.

